# Indications that Stockholm has reached herd immunity, given limited restrictions, against several variants of SARS-CoV-2

**DOI:** 10.1101/2021.07.07.21260167

**Authors:** Marcus Carlsson, Cecilia Söderberg-Nauclér

## Abstract

“When COVID-19 cases go up, public compliance with restrictions is poor, when cases go down, public compliance is good.” In this article, we question this explanation and show that relatively low levels of sero-prevalence helps to keep cases down. In other words, the herd-immunity threshold appears to be much lower than previously thought. We construct a mathematical model taking pre-immunity, antibody waning and more infectious variants of concern into consideration, thereby providing a theoretical framework in which the cases in Stockholm county can be fully predicted without relying on neither oscillations in restrictions (and public compliance thereof) nor vaccination roll-out. We also show that it is very difficult to match the data from Stockholm without including pre-immunity, or, which turns out to be equivalent, great variations in susceptibility.

## 1 Herd immunity at 22.6% sero-prevalence?

In march 2020, when it became clear that the COVID-19 outbreak was turning into a global pandemic, mathematical models had a huge impact on public policy. For example, the imperial college report no 9 predicted 500000 deaths in UK in the worst case scenario [11], given that the government did nothing to mitigate or suppress the pandemic, which led to the subsequent strategy shift towards suppression and eventually lock-down on March 23rd. The Swedish government on the other hand decided to keep society relatively open and have maintained limited and rather fixed NPI’s throughout the pandemic, making Sweden an excellent example on which to test the validity of such mathematical models. In this study, we use the metropolitan area of Stockholm with about 2400000 inhabitants to test how mathematical models and real outcome correlate in reality.

We consider the time-series of COVID-19 positive PCR-cases in Stockholm county since 1st September 2020, as displayed in Figure 1. Two distinct waves are clearly visible, which we will refer to as the second and third wave, given that the first wave of spring 2020 is not included. Before the second wave hit, the sero-prevalence was around 10% in Stockholm county, which rose to 22.6% after the second wave but before the third wave was initiated [14] (sero-prevalence data obtained from blood-donors), which by extrapolation gives a sero-prevalence in Stockholm of about 33% in the end of May 2021. Adding to this, in late April when the third wave began to bend downwards, about 5% of the population were fully vaccinated and 13% had received their first shot. In the age group 0-69, the vaccine coverage was practically zero apart from hospital workers and people from other selected professions.

**Figure 1:**
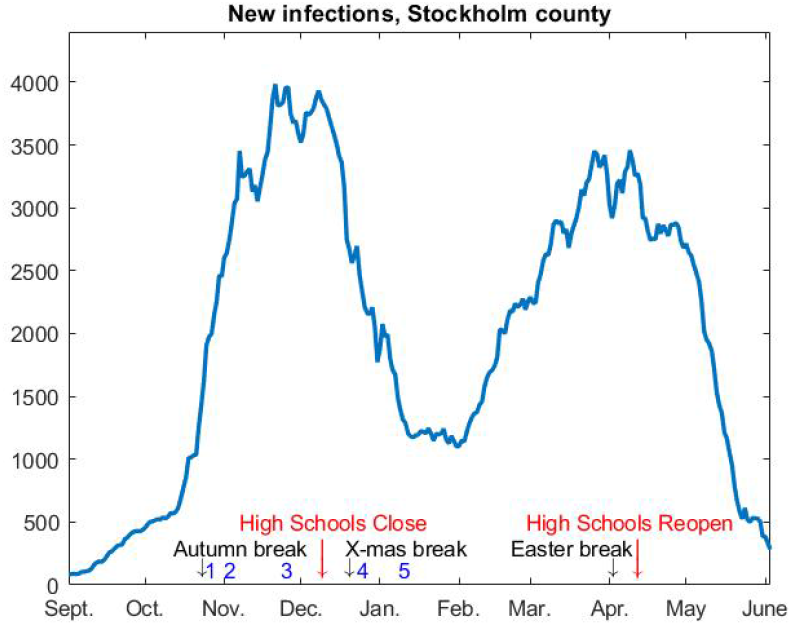
7-day rolling average of new cases in Stockholm county, adjusted to match the current sero-prevalence.

Given these figures, most scientists observing this time series would conclude that NPI’s along with voluntary behavioral changes in the population made the second wave bend downwards in early November, and then guess that public weariness and/or mutant strains caused the third wave, which again bent downwards due to renewed public compliance with recommendations in the face of the recent surge. Indeed, this is the by now accepted scientific consensus among scholars studying the pandemic, which is sometimes called “herd-protection”, and builds on the simple idea that when a major deadly epidemic hits, society reacts in a way that is impossible to predict mathematically. Therefore, mathematical models are often grossly wrong, as exemplified by previous potential disasters, such as various Ebola outbreaks [2, 8]. However, in order for this to be the key explaining factor for the trajectory in e.g. Fig.1, large fluctuations in the effective *R*-value, denoted *R*_*e*_, are needed. This is seen for example in Figure S1 in [17], which focuses on modeling Manaus with a SEIRD equation system with variable *R*_*e*_, where *R*_*e*_(*t*) oscillates between 0.8 and 4 in order to get a good fit. The pandemic response in Sweden challenges this interpretation, and in this article we explore an alternative, and in our opinion a more plausible, explanation for the viral spread pattern in society.

About a year ago, a number of different researchers put forward the idea that pre-immunity or immunological “dark matter” could lurk behind the unexpected unfolding of the COVID-19 pandemic, but as time went on this dark matter was not found and along with various erroneous predictions involving pre-immunity [15], the hypothesis seems to have been discarded. This could also be related to a fear of causing unnecessary deaths related to political implications of such scientific findings, connected with the Great Barrington Declaration and a small group of scientists pushing for the removal of NPI’s and mitigation strategies [19]. If a level of pre-immunity would exist it would give fuel to that flank of the “scientopolitical discourse”.

Be that as it may, we believe that it is too early to discard the hypothesis that some sort of pre-immunity needs to be taken into account, in particular for accurate mathematical modeling. In a parallel publication [7] we will demonstrate that what looks like pre-immunity on a population level, could in fact be a consequence of large variability in susceptibility on an individual level, which in turn could depend on innate immunity, cross-reactive protective immunity initiated by another virus and a number of other factors. We speculate that, given the current Swedish restrictions, the dosage that most people are exposed to in society is not strong enough to infect those who have a low susceptibility to infection with SARS-CoV-2, and hence they appear as immune despite not having had COVID-19. We also show that mathematical models taking this complex structure of variable susceptibility to SARS-CoV-2 (as well as super-spreaders) into account is, from a practical point of view, equivalent to simpler models that incorporate pre-immunity. Therefore, “pre-immunity” should not necessarily be interpreted as that certain people have sterilizing immunity due to a single factor. We discuss these aspects in greater depth in [7], and focus here on showing that pre-immunity is a necessity for successful mathematical modeling of the pandemic. To avoid confusion with sterilizing pre-immunity, we will occasionally refer to it as “population pre-immunity”. We argue that this is the key factor that has protected Sweden from a much higher hospitalization rate and death toll, given the Swedish mitigation strategy, and that it helps to keep cases down to a much greater extent than predicted by traditional models for disease spread.

Since practically all high-resource countries in the world opted for severe and variable NPI’s for the last 12 months, and thanks to the arrival of vaccines, it is quite likely that we would never have found out whether a population pre-immunity existed or not. If it wasn’t for Sweden which, to everyones surprise, decided to combat the pandemic with relaxed NPI’s that have been rather fixed, and only minor modifications were implemented when cases went up or down.

If, as we argue here, the Stockholm cases have plummeted twice due to depletion of susceptibles linked to pre-immunity, rather than variation to the NPI’s and public compliance thereof, it is a question of semantics whether to call it herd-immunity or something else. On one hand, cases can still go up if NPI’s are lifted, so the term herd-immunity may be misleading. In fact, anytime cases plummet in any country, one could argue that they have reached herd-immunity with their present NPI’s, which would be misleading. The point is that the restrictions in Sweden are very relaxed and that society is functioning practically as normal, with banning of major gatherings and bars/restaurants closing early as the only main limitations, and a large part of the population is acting as if the virus did’t exist. We call it herd-immunity under limited restrictions.

## 2 The Swedish pandemic response

From a mathematical modeling perspective, Sweden provided a unique opportunity to study the be or not be of this hypothetical pre-immunity, in particular the densely populated metropolitan area of Stockholm with about 2.4 million inhabitants. The only noteworthy change in NPI’s during the time-frame Sept 2020 to May 2021 was the closing of high-schools (age-group 15-18 years) on December 7, 2020 and subsequent reopening on April 1, 2021. Bars and restaurants have been open but only with table-service and a maximum of 4 people at each table. In addition, the following changes in the recommendations were made (labeled 1-4 on the timeline in Fig. 1) 1: Recommendations for elderly (70+) to self-isolate are removed and replaced by same recommendations as the general population. 2: Before November 1st the maximum amount of visitors to a public event was 50, which then was changed to 300, although a cap on maximum 50 participants in dancing events remained. 3: Prohibited public gatherings involving more than 8 people (shopping, restaurants, bars etc. were exempt from this rule). 4: Alcohol-sales only until 20.00, compared with 22.00 previously. 5: Facemask recommendation is issued for health care visits and rush-hour public transport. This was followed by low public adherence, less than one out of three was reported to follow the recommendation based on random testing performed by the Swedish Public Broadcasting company [20]. Note that the *relaxations* 1-2 coincide with the time when the second wave looses momentum and levels out, in opposition to what one would expect.

Neither these policy-updates nor any of the major holidays (Autumn, Christmas, Sport- and Easter-school holidays^1^, see Figure 2) had any notable effect on the two waves. Besides NPI’s, it is of course possible that the population itself isolates whenever infection transmission rates are high, which could cause the ups and downs in cases (known as herd-protection [17]). However, Figure 2 displays Google-mobility data (i.e. in which type of location people spend their time) for Stockholm county, and it is very hard to see any correlation between the fluctuations and the epidemic waves in Figure 1. This leaves the possibility that major fluctuations in *R*_*e*_ are caused by people taking precautions that are not seen on Google mobility data, such as keeping sufficient distance to others in work places/public transport/stores etc. However, anyone who has tried to avoid people in a public place, that are not concerned with avoiding you, understands that this explanation is unlikely. Moreover, apart from the difficulty of doing this in practice, one has to bear in mind that the Swedish population has been informed by the Public Health Agency (PHA) that frequent washing of hands and keeping 2 meter distance between people are the key factors to limit the spread of the virus in society. As SARS-CoV-2 is an airborn transmissible virus, found in the air surrounding infective people [16], these recommendations are expected to have had a limited value to protect people from becoming infected. Furthermore, aerosol spread is continuously denied by the PHA as an important source of infections, despite both CDC and WHO recently changing their stance, which for example has led to that good ventilation has not been part of the pandemic preventive measures. Less than one third of the population use voluntary protective gear, such as face masks, and among those who did try to protect themselves, face-shields were about as common as face-masks, due to the wide-spread misconception that SARS-CoV-2 primarily spreads by infected individuals coughing and sneezing, which has been repeated over and over by spokesmen for the PHA. Since COVID-19 can spread by aerosols [16], particularly in indoor climate, we argue that the conditions for steady spread of COVID-19 were good throughout the time-period studied, in particular the cold and rainy months between October 2020 and May 2021 (April and May 2021 were unusually cold).

**Figure 2:**
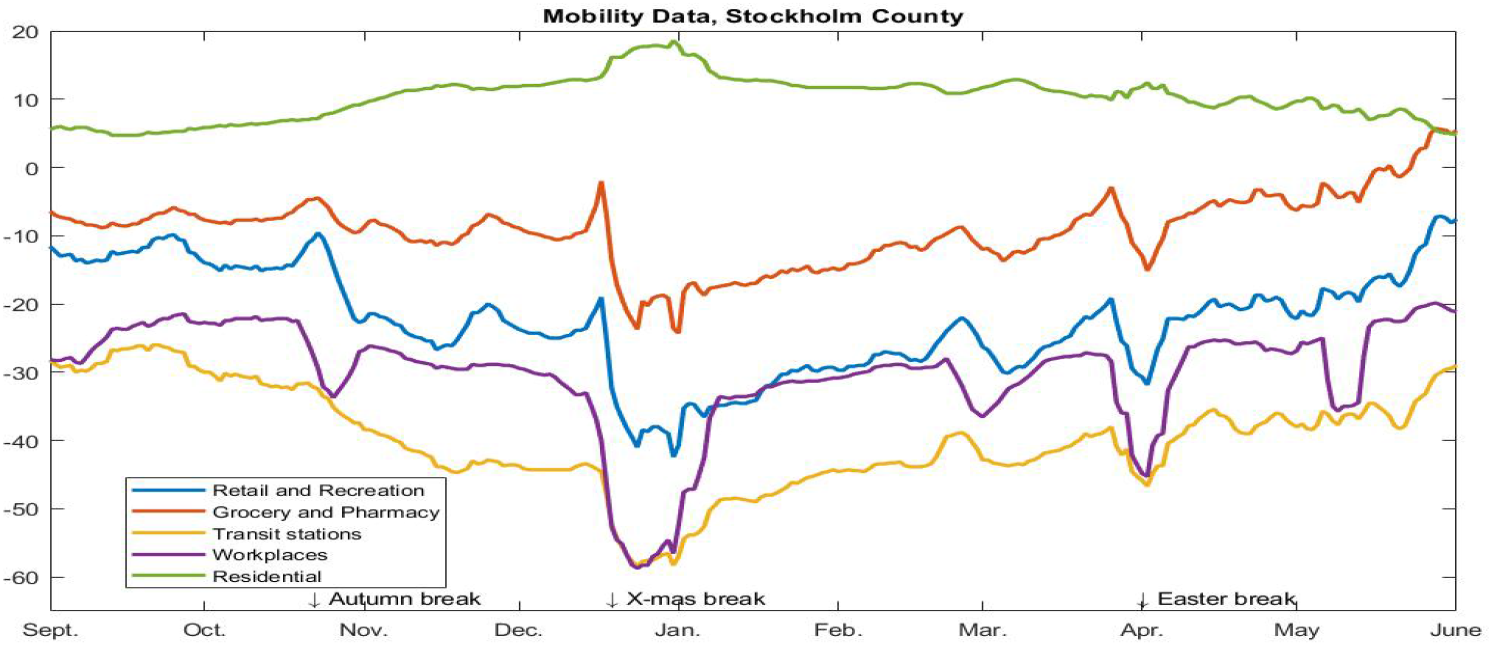
Google-mobility data seems to have no bearing on the COVID-19 curves in Figure 1.

These observations led us to search for alternative explanations to understand the dynamics of virus spread seen in Figure 1. We have three parallel studies on the topic of pre-immunity [6, 7, 24]. The first provides a profound review of different ways to model COVID-19, trying to mathematically explain the unexpected unfolding of the pandemic in Stockholm county in the absence of population pre-immunity, but ultimately failing, leading to the conclusion that a population pre-immunity of around 60% seems to be present in the Swedish population. The second publication shows mathematically that what is perceived as pre-immunity on a macroscopic level may be a manifestation of variability in susceptibility on a microscopic level (hence the term *mathematical* pre-immunity). In the the third study we show that the variations in susceptibility could be linked to a cross-protective immunity between Influenza A strains, implying that previous influenza infections and/or vaccinations could provide a level of protection against SARS-CoV-2.

## 3 Mathematical modeling

In this section we describe the key features of our mathematical model, which is an extension of a classical age-stratified SEIR-model. The key novelties are the incorporation of susceptibility variation via a pre-immunity parameter *θ* and variants of concern. For accuracy we also include the waning of antibodies and vaccinations. Besides *θ*, the other key parameter is *β* (transmission rate), which is related to *R*_0_ via the formula

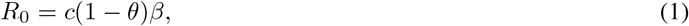

where *c* is a constant (see (7) for details). To a first approximation, it holds that the amount of newly infected at time *t* is given by multiplying the amount of infectives with the number

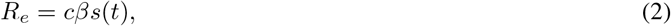

where *s* the fraction of susceptibles, with initial value *s*(0) = 1 − *θ* (more accurate formulas are provided in the supplementary material, Section 5.3). In mathematical language, the key question of this paper is whether the transmission rate parameter *β* is constant or whether it too should depend on time. Since we multiply with *s*(*t*), the *R*_*e*_-value gradually decrease as susceptibles become fewer, and the herd immunity threshold is reached when *R*_*e*_ = 1. The key insight of [6] is that we can get an almost perfect fit with a fixed *β* if we include a high value of *θ*, above 0.6. In the experimental section of this paper we will try both alternatives, the other being *θ* = 0 and time-varying *β*. The key difference between the the two explanations is that in the former, the transmission rate parameter *β* will be much higher and removing a small amount from the susceptible group, say 20%, can make a substantial relative change in *s*(*t*), thereby reaching *R*_*e*_ = 1 at much lower values of sero-prevalence. To be concrete, if *R*_0_ = 1.5 and *θ* = 0.5, then an increase of sero-prevalence of 20% gives 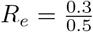, whereas in the latter scenario we would get *R*_*e*_ = 0.8 · 1.5 = 1.2. First, we briefly describe the new features of our updated model, the mathematical details of which is presented in the Supplementary Material (SM).

### 3.1 Anti-body waning

While most would agree that antibodies wane, there is no consensus on how fast that happens. In a study [5] by Buss et. al., the half-life of SARS-CoV-2 antibodies was estimated to 5 months, which in part could explain the second wave in Manaus. However, this estimate is at odds with other key studies such as [9], which established a “moderate” decline of anti-bodies over 8 months. Moreover, the 5 months estimate seems to be at odds with the subsequent publications from the team behind [5], which estimates the reinfection rate to be at most 25% [22]. Given the acclaimed 76% attack rate of the first wave and the fact that the second wave was larger, this means that by now over 150% must have had COVID-19, which implies a much higher reinfection rate or a significantly increased Infection Fatality Ratio (IFR). Both explanations appear unlikely.

This is not the only problematic issue with the numbers reported in [5], which has been discussed in [18] and carefully examined from a mathematical perspective. The measured sero-prevalence of 44% in Manaus in June quickly dropped to around 25% in October, which implies a very strange profile for the waning of anti-bodies to add up. A more likely explanation for the measurement of 44% in the midst of a raging first wave would be that only “high-risk” individuals and people having passed the infection, would dare to give blood under such circumstances, as argued in [18]. These authors then go on to hypothesize that the maximum sero-prevalence in the population more likely was below 30%, which would make the anti-body waning pattern more in line with other studies, as well as put the IFR in Manaus more aligned with IFR’s observed elsewhere in Brazil. An alternative explanation offered by the authors of [5] is that IFR of the Brasilian variant P.1 is substantially higher. This would mean that the second wave in terms of cases was much smaller than the first, despite larger amount of deceased, which seems rather unlikely.

Summing up, it seems more logical to assume that the half-life of antibody waning is much more than 5 months, but nobody knows the exact figure. In this study we are not aiming to predict the future with precision in a long term perspective, so we proceed with an estimate of 1.5 years that fits well with the Swedish data from [14]. More precisely, Stockholm had 11.3% sero-prevalence in early June and 9.6% in mid October (which most likely is unaffected by the rise in cases seen in Figure 1, taking into account that sero-conversion takes a few weeks). With a half-time of 1.5 years this yields a perfect fit and puts the antibody level on the first of September in Stockholm (the starting date for our modeling) at 10%. In the end, the modeling in short term is fairly unaffected by the exact rate of antibody waning, as it mainly affects the long term behavior, which will be more dependent on vaccination schemes than anti-body waning. Still, it is interesting to include this parameter in the model, partially to get the numbers as accurate as possible, and partially to see the long term development in the absence of vaccines (or, for that matter, in the scenario that new variants keep getting ahead of vaccines).

### 3.2 Mathematical pre-immunity

A fair argument against the hypothesis of pre-immunity is recurring reports of quire rehearsals and similar events, where a majority get infected at one super-spreading occasion. To counter this argument, we first note that singing produces several orders of magnitude more aerosols than normal talking [1], and that SARS-CoV-2 is spread via aerosols [16]. Hence, participants to such events are likely to get exposed to a very high dosage in case a super-spreader is present. Secondly, pre-immunity, or immunity past infection as well for that matter, is not a binary variable that either gives a 100% protection or zero. One of the take home messages of [6] is that there is practically no difference mathematically between an advanced model taking variable susceptibility into account, and a more naive model which simply stipulates that a certain proportion of the population is pre-immune. Hence, the idea of a binary pre-immunity works well on a macro-level as long as one is aware that this is a simplification that does not always apply on a micro-level, where vocal activity, ventilation, face-masks and a number of other factors could make a group more or less prone to becoming infected in larger numbers.

Another take-home message of [6] is that various versions of SEIR-models, including variable infectivity, activity levels and socio-economic factors, have a marginal effect on the key characteristics of the corresponding curves, as long as the corresponding parameter values are kept within reasonable limits. Adding one or several of the above factors typically results in output that deviate 10% to at most 20%, far less than needed to explain observed data. Based on this, we find that a simple SEIR-model with age-stratification, developed by Britton et. al. [4] (with a “contact matrix” adopted to Swedish age distribution, see [6] for details), in combination with a binary population pre-immunity is the best choice for accurate modeling of real data. We also updated the contact matrix to take into account isolation of the elderly, and calibrated it to match Swedish incidence of COVID-19 in the various age groups, see SM 5.2 for details.

In this study we run our model with a pre-existing immunity of 62% for Stockholm, which we base on a new mathematical method which estimates the pre-immunity directly from a time series of cases and available sero-prevalence information [24]. Applying this to the Stockholm county second wave (red curve in Figure 3), gave an immunity level of 72%, which would correspond to a pre-pandemic immunity of 62% (upon subtracting the 10% having developed COVID-19 anti-bodies by September 2020).

**Figure 3:**
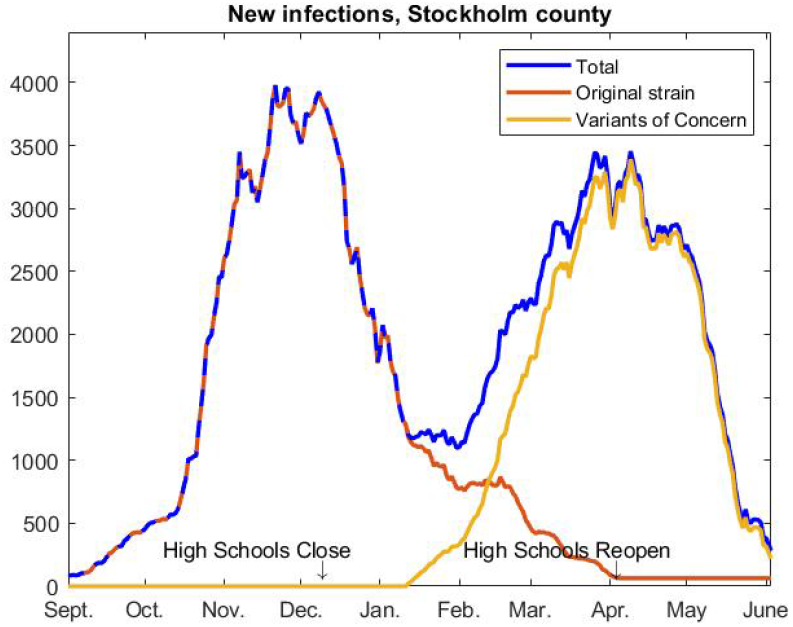
Total cases separated in original strain and variants of concern (almost exclusively due to alpha). Data is not available from April and onwards, hence the straight line at the end of the red curve.

### 3.3 Variants of concern

In order to accurately model the pandemic in Stockholm, we need to separate the second from the third wave, and more importantly understand what caused the latter. Based on publicly available data from the Swedish Public Health Agency [13], it follows that the third wave in Stockholm is entirely caused by the “British” Variant of Concern (VoC) B.1.1.7, now referred to as alpha. The other VoC’s, P.1, B.1.167 and B.1.351, seem to have receded and at most caused 6% of the cases (in the month of February). To better see the dynamics behind the two waves in Figure 1, we have separated the cases caused by VoC’s from the rest, which we for simplicity will refer to as the “original strain” of SARS-CoV-2. In Figure 3, we display cases caused by the original strain and by VoC’s independently. This suggest that the epidemic in Stockholm county would have been over by mid-April, this would mean that Stockholm should have reached herd-immunity (given limited restrictions) around mid-April, at a sero-prevalence of around 27%.

Of course, it is tempting to add up 27% with the pre-immunity of 62% and say that the total immunity was 89%, which is in line with expectations for the real Herd Immunity Threshold (HIT). This would be perfectly in order, if it turns out that there is a single cause factor explaining the 62% pre-immune, such as a cross-reactive antibody-protection found in 62% of the general population. As mentioned earlier, we provide evidence that previous Influenza A infections could have provided such high protection rate against SARS-CoV-2. However, we believe that this manifests itself in large variability of susceptibility to SARS-CoV-2, and not sterilizing immunity. Confirming this hypothesis will require further studies, as other viruses may also mediate a protective pre-immunity. Furthermore, genetic factors may also provide different protection on an individual level due to differences in the innate and adaptive immune response against this virus. In particular, the 62% could easily drop to some lower value if people start exposing themselves to higher risk or if a virus variant appears which has the ability to circumvent the mechanisms behind this phenomenon. Indeed, the data from India seems to suggest that pre-immunity to delta is significantly lower. In summary, we prefer to not sum up the two immunity variables since it may give a false illusion of a high degree of protection in society.

In order to model the third wave, we need to update the parameters for the new strain alpha. As shown in [7], section 4.2, both transmission rate *β* and pre-immunity *θ* need to be updated. In order to do so, one needs a time series from a major outbreak, it is presently not possible to determine these constants theoretically. In other words, we will pick model parameters so that the outbreak of the VoC’s, which is modeled separately, fit well with the yellow curve.

### 3.4 Vaccination

The first vaccinations started in week 52, and it was not until early April that people younger than 70+ started to get vaccinated (with the exception of special work groups such as health care workers). We remark that it is hardly the elderly that “drive the epidemic”, and moreover it takes quite some time from the first shot to maximal protection attained after the second shot. These observations alone suffice to prove that it was not the vaccination schemes that led to the turn-around of the third wave in early April, seen in Figure 3 (and most definitely not the second). However, to be on the safe side, we have included vaccine roll-out in our model and (generously) estimated that people develop 90% protection within 3 weeks of the first shot. We do this to demonstrate that, even with an exaggerated help from vaccination, the common explanation of COVID-19 dynamics which only focus on variable *β* due to NPI’s etc., is not very convincing. The graph of vaccination rollout is shown in Figure 6.

## 4 Results

We first show what our mathematical model implies for Stockholm County in the short and long perspective, then we compare the results with an alternative model that does not involve any pre-immunity, but instead rely on changes in NPI’s and public behavior in order for the model and reality to match. While these are not mutually exclusive, we shall see that the former model gives more realistic output, showing that “herd-protection” alone is an unlikely explanation of what happened in Stockholm county.

### 4.1 Stockholm, with population pre-immunity *θ*

The model in Figure 4 uses a 62% population pre-immunity against the original strain and a 52% population pre-immunity against alpha, combined with a slightly higher transmission parameter *β*. We also, at the start of the model, put 10% in the recovered group since this was the sero-prevalence (of anti-bodies caused by infection with SARS-CoV) present in early September, before the start of the second wave.

**Figure 4:**
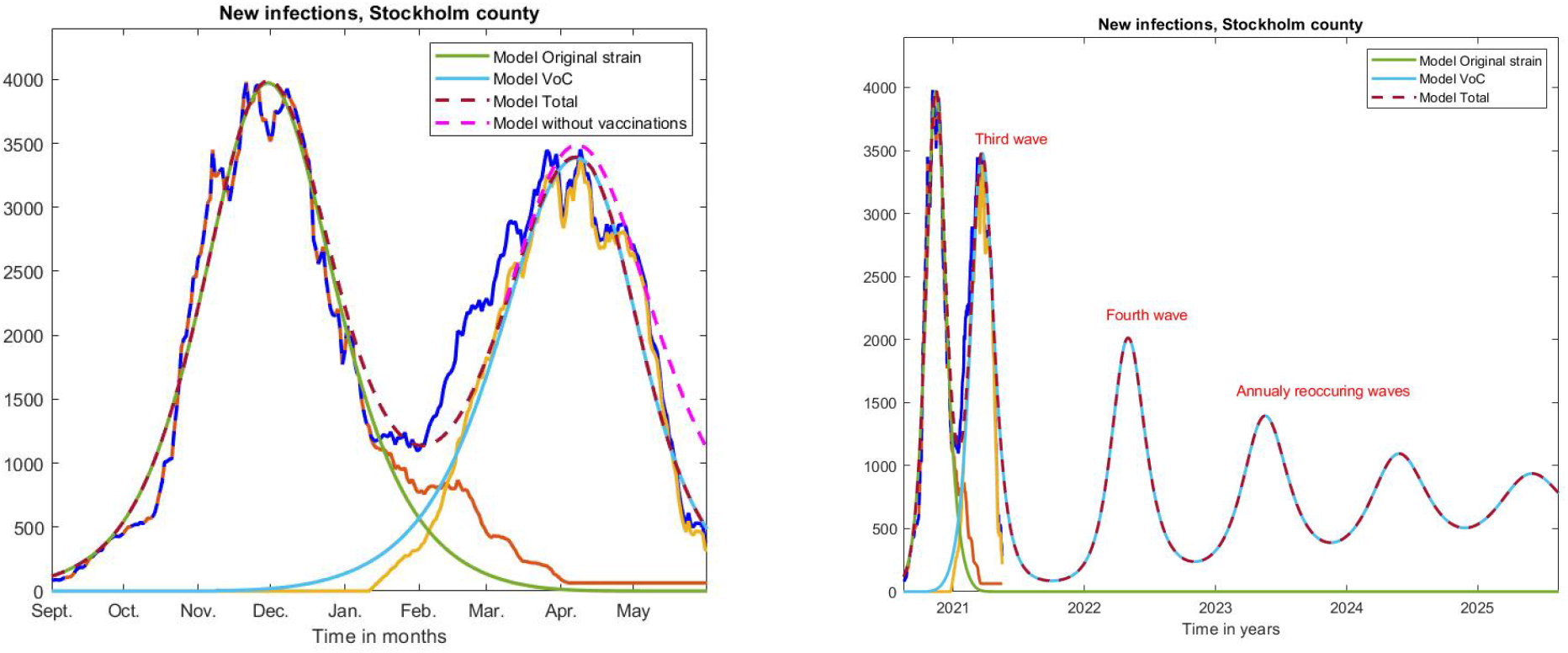
Left; same curves as in Figure 3 on a longer time-scale, along with curves produced by our model.

The result of running the model is seen in Figure 4 (left). Green is the color for the original strain and light blue represents the variants of concern. As is plain to see, the model gives a very accurate fit, with sections of the measured curve being partially hidden model curves. Of course, it is easier to model the past than predict the future, so therefore we would like to underline that the curve of total cases seen in the above model was published on Twitter as a prediction in early May, before the fast decline of new cases.

For the second wave (green) we chose a transmission rate parameter *β* = 0.38, corresponding to *R*_0_ ≈ 1.4 for the “original strain” in the beginning of the second wave. To get a good fit for the third wave, we adjust the population pre-immunity down from 62 to 52 percent, while increasing transmission rate *β* only slightly to 0.38. This gives an *R*_0_ for alpha of around 1.5. Note that this is perfectly compatible with reports of alpha being 40-100% more infectious, since a division of the effective *R*_*e*_-values for the original strain and the VoC’s on the first of February gives values of around 1.75.

Here is an important point speaking in favor of the underlying interpretation behind the above model, which relies on a “population pre-immunity”. The original strain was estimated to have an *R*_0_-value above 2, then alpha was estimated to be around 43-90% more infectious [10], and now reports show that the delta strain is almost twice as infectious as the alpha strain. But then the *R*_0_-value of delta should be above 6, which is highly unlikely. With such a high *R*_0_−value, delta should infect the majority of a fully susceptible population within a matter of 2 weeks, whereas it took two months to reach peak in India. With the interpretation above, the VoC’s are not necessarily having a much higher transmission rate, but rather can infect a larger proportion of the population, making it seem more infectious in the initial phase.

If this model/interpretation is correct, then Stockholm practically has reached herd-immunity under limited restrictions and no further major outbreaks are to be expected, unless recommendations and behavior change drastically. In fact, if it hadn’t been for alpha, it seems that the herd-immunity under limited restrictions was first reached in early December, just before the closing of high schools. This is because, if no updates are done to the NPI’s, the herd immunity threshold is reached shortly after the peak of the wave, not after it has receded (see SM Section 5.3 for a fuller discussion of this topic). This is the reason why a major outbreak usually ends with a final size substantially larger than the Herd Immunity Threshold. By the same logic, the HIT for alpha was reached in mid April (14th of April to be precise), when most people in the group 0-69 were still unvaccinated. In the graph we also plot the model output without vaccination scheme, indicating that the HIT would have been reached at the same time anyways. The main effect of the vaccinations seems to be a faster decline of the spread, as well as significantly fewer hospitalizations and deaths (not shown). Other examples of places that reached herd-immunity under restrictions and then were hit by major new waves include Manaus in Brazil and India. It is clear by these observations that the herd-immunity threshold, with or without restrictions, is rather unstable and can at any time be moved upwards due to new variants. Hence, it seems to be a bad policy to rely on it.

A weakness of this *hypothesis* is that it is not clear which factor or combination of factors that causes the pre-immunity, and it is certainly not clear whether pre-immunity is long-lasting or whether it also wanes over time, which changes the modeling in the longer perspective. Nevertheless, it is interesting to see how the model behaves in the long run, in the absence of both vaccinations and future mutations. Figure 4 (right) displays the result. As is plain to see, recurring waves become milder and milder to finally level out at a constant rate of around 700 new infections per day, equalling 29 cases per 100000 per day. Put differently, this means that every person will get COVID-19 with cycles of about 10 years. Another interesting observation is that alpha completely takes over and the “original strain” never comes back. It is likely that the Indian variant delta will have the same effect on alpha in Sweden and generate a new wave during the fall, as seems to be the case in Great Britain at the time of writing. This exposes the weaknesses of mathematical modeling for predictions since it is impossible a priori to know which values of transmission rate *β* and pre-immunity *θ* will work for a new strain. However, this uncertainty is still far less than the uncertainty provided when modeling with zero pre-immunity and variable *β*. We now take a look at this option.

### 4.2 Stockholm, with variable transmission rate *β*

Without going into details, which are found in SM Section 5.5, in order to model with variable transmission rate due to changes in behavior/NPI’s, we replace (5) by

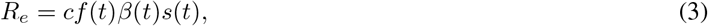

where now *β*(*t*) is a function that is initially fixed at the transmission rate *β*_*OS*_ of the original strain, and then slowly increases to a new higher level *β*_*V oC*_ corresponding to the transmission rate of the VoC’s (i.e. alpha in the case of Stockholm). The function *f* (*t*) incorporates the changes in behavior/NPI’s, in such a way that we get an exact fit with the observed cases seen in Figures 3 and 4. We calibrate the choice of the original transmission rate *β*_*OS*_ so that the average of *f* in September equals one. At this point in time, most people were convinced there would be no more major waves in Sweden, and the PHA and chief epidemiologist downplayed the situation and described a 35% increase as “a small cautious increase” [23]. Shortly thereafter a second and later a third wave hit Stockholm. We therefore argue that peoples behavior in September was close to normal, so *f* (*t*) ≈ 1 means that people take minimal precautions, given the NPI’s in place.

Next issue is how to pick the value of the higher transmission rate of alpha. The increase in transmission rate was measured to 56% in England [10], and this corresponds precisely to the increase in *R*_*e*_ as given by running EpiEstim online *R*-estimator [25]. However, note that a higher value of *β*_*V oC*_ leads to a lower value of *f*, and vice versa. To remove any doubts that our model is somehow constructed to favor the conclusion we wish to make, i.e. that changes in behavior and NPI’s can not be the full explanation behind the unexpected curves in Figure 3, we have decided to make *β*_*V oC*_ only 38% more transmissive, which is the lower end of the 95%-confidence interval estimated in [10]. We thus set *β*_*V oC*_ = 1.38*β*_*OS*_.

The result of running the model on the Stockholm cases is seen in Figure 5, (left), where the graph of total cases from Figure 3 has been added below for easy comparison. It is clear that in order for the second wave to turn around, as it did in November-December, we need a reduction in transmission of around 20-30% based on behavioral changes. Then a slight relaxation is seen at the same time as the arrival of alpha, although this is an effect of the fact that we chose a very modest transmission rate *β*_*V oC*_. Still, in order for the third wave to bend down, as it does in April, we need a reduction of 35% in *f*, followed by a reduction down to levels below 50% to cause the sharp decline in May.

**Figure 5:**
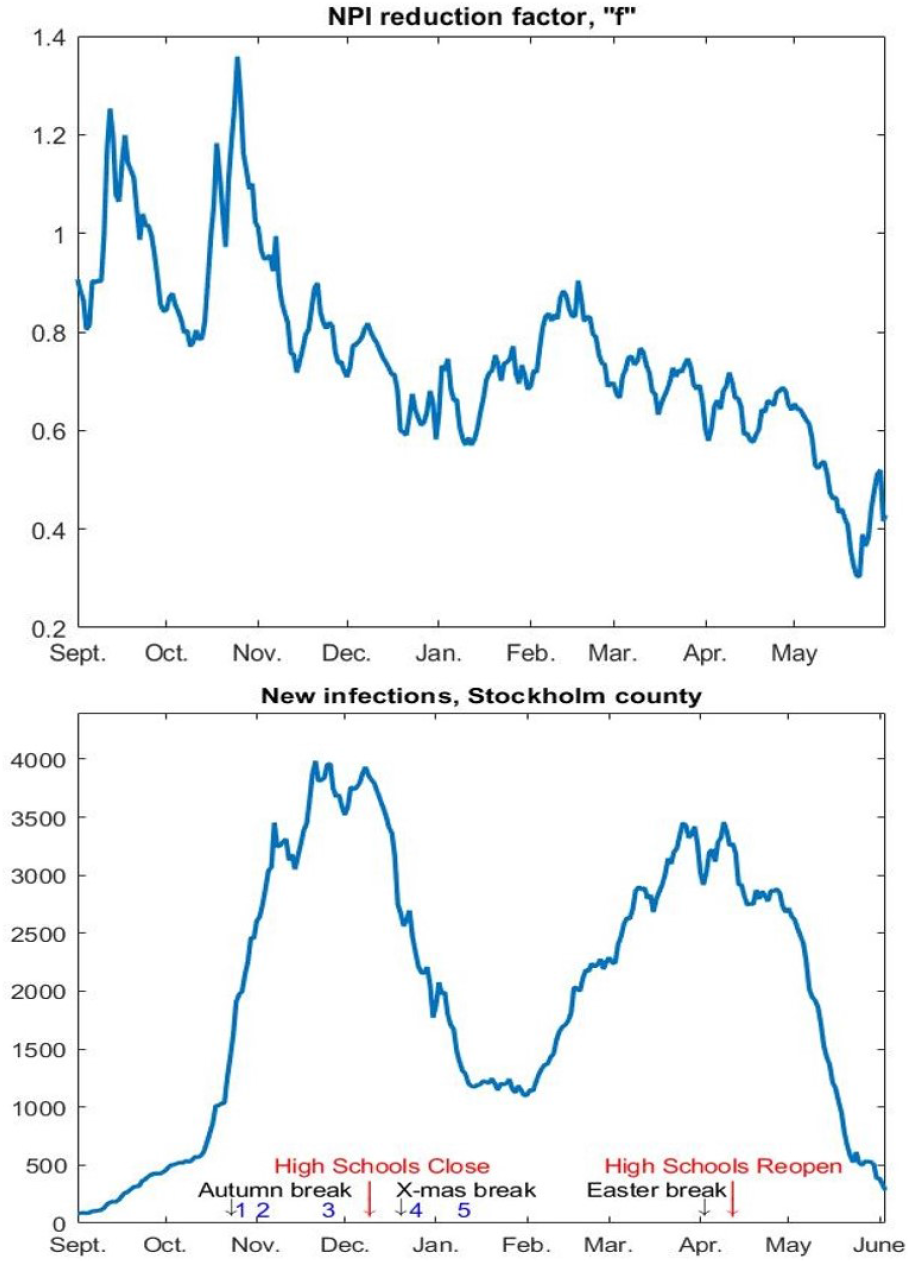
Variations in behavior/NPI’s in order to get a perfect model fit for Stockholm.

For us who have lived in Sweden during the pandemic, the above seem to be an unlikely scenario. While this is not an entirely scientific punch-line, we remind the reader that the model already takes into account a substantial isolation of the elderly, and that we have chosen both *β*(*t*) and vaccination effects to keep *f* as high as possible, so the graph in Figure 5 is more like an upper bound. Of course, one could also argue that the final drop seen in May is caused partially by better weather, (despite May being unusually cold). Although it is unclear exactly to what extent this reduces transmission, it undoubtedly does have an impact. However, by the same token we should then increase the transmission rate after September, which also had warm sunny weather. This would lead to an overall reduction of *f* throughout the period, and hence even less probable output.

## 5 Discussion

This work challenges the established interpretation of variable NPI’s and “herd-protection” as the *main cause* for the rise and fall of epidemic waves in *hard-hit* locations where authorities have been either unwilling or incapable of enforcing strict NPI’s, such as Stockholm. If we assume that changes in behavior among the population had a marginal effect on the spread in Stockholm, we show that herd-immunity (given the limited restrictions) has been reached twice, first in early December against the original strain, and then in mid April against the alpha strain, and that this happened more or less independent of updates in the NPI’s and the vaccination roll-out. Consequently, our work indicates that herd-immunity thresholds are much lower than traditional mathematical models suggest, but on the other hand that such thresholds are unstable and will move upwards upon introduction of new variants of concern. A theory for the underlying mechanisms of why the herd-immunity threshold is lower than generally believed, which evolves around the idea that variable susceptibility to SARS-CoV-2 significantly alters corresponding formulae, is put forth in the adjacent paper [7]

These results of course do not imply that NPI’s and behavioral changes do not alter transmission rates, so while we here run our models with a fixed value of transmission rate, in reality it should vary on both NPI’s and other factors such as season. In other words, we do not argue against NPI’s, we simply argue that it is difficult to envision that they alone can explain the outbreaks seen in Stockholm County during the second and third waves.

It is therefore important to stress is that this work by no means implies that it is safe to lift NPI’s, on the contrary, it implies that around 60% could have some protection against SARS-CoV-2 under current NPI-levels. Such protection could disappear due to e.g. unfortunate mutations, protection-waning, as well as exposure to higher viral doses following careless lifting of restrictions. Adding to this, if a pre-existing cross-reactive immunity does exist then it does not imply that NPI’s and voluntary public changes in behavior do not affect at all, and hence the true value of individuals with pre-immunity could be substantially lower. Finally, on an individual level it is impossible to know if pre-immunity is present or not, and it is well established that getting COVID-19 unvaccinated is connected with greater health risks than if vaccinated. Based on this, it is our firm conclusion that the vaccination roll-out must continue with high participation to avoid both personal tragedies and COVID-19 becoming endemic, as seen in Figure 4. On the other hand, since all NPI’s are associated with costs and other risks, we hope this work can help to establish better prediction models to support difficult decisions.

## Supporting information

Supplementary Material: Mathematical details

## Data Availability

All modeling relies on publicly available data from the Swedish Public Health Agency

## Footnotes

1 Autumn-holiday, week 44, Sport-holiday week 9, Easter holiday week 14.

