## Supplementary Material: Mathematical details for "Indications that Stockholm has reached herd immunity, given limited restrictions, against several variants of SARS-CoV-2"

### 5.1 Age-stratified SEIR with antibody waning

We briefly recapitulate the age-stratified SEIR model developed by Britton et al [4], which is described in greater detail in the supplementary material of [6].

We divide the population in 6 age-groups and let $\mathbf{s}, \mathbf{e}, \mathbf{i}$ and $\mathbf{r}$ represent column-vectors including the fraction of the full population in respective age-group of the classes Susceptible, Exposed, Infective and Recovered. A contact matrix $A$ determines the amount of contacts between the groups and a coefficient $\beta$ the rate of transmission. The heart of the equation system is the term

$$\beta \text{diag}(\mathbf{s}(t)) A \mathbf{i}(t) \tag{4}$$

which determines the rate of new cases in each respective group, but this needs to be slightly modified for the following reason; Adding pre-immunity is most easily done by initializing $\mathbf{s}$ by $\mathbf{s}(0) = (1-\theta)\mathbf{w}$ where $\theta$ is the fraction of pre-immunity and $\mathbf{w}$ the vector with the fraction of the population in the respective age-group. However, as explained in [7], the pre-immunity $\theta$ (which we have estimated to 0.62 for Stockholm) is most likely a modeling artefact of the phenomenon that susceptibility to the virus is very variable between individuals. A plausible assumption is that this changes very slowly over time, so therefore this number stays fixed during the modeling. However, upon introducing a variant of concern, we obtain a new value of $\beta$ but potentially also a new value for $\theta$ (see [7], Section 4.2, for a fuller discussion of this topic). Since we can not suddenly change the initial value in the model, we instead opt for setting $\mathbf{s}(0) = (1-\psi)\mathbf{w}$, where $\psi$ is the level of acquired immunity $\psi$, e.g. $0.1$ in the examples for Stockholm. This way the whole remaining population is technically "susceptible", but the pre-immunity $\theta$ can be incorporated by replacing the term (4) by

$$\boldsymbol{\nu}(t) = \beta \text{diag}(\mathbf{s}(t) - \theta \mathbf{w}) A \mathbf{i}(t). \tag{5}$$

This way the total amount of susceptibles will never exceed $1-\theta$, but this limit can be moved upwards by adjusting $\theta$ for more aggressive variants. As long as $\theta$ is held fixed, it is easy to see that the two alternatives are equivalent in terms of the effect on the remaining functions $\mathbf{e}, \mathbf{i}$ and $\mathbf{r}$. These relate to $\mathbf{s}$ as follows

$$\begin{cases} \mathbf{s}'(t) = -\boldsymbol{\nu}(t) + \tau \mathbf{r}(t) \\ \mathbf{e}'(t) = \boldsymbol{\nu}(t) - \sigma \mathbf{e}(t) \\ \mathbf{i}'(t) = \sigma \mathbf{e}(t) - \gamma \mathbf{i}(t) \\ \mathbf{r}'(t) = \gamma \mathbf{i}(t) - \tau \mathbf{r}(t) \end{cases} \tag{6}$$

where the main new feature is the term $\tau \mathbf{r}(t)$ which takes the waning of antibodies into account. Since we have decided to put half-life to 1.5 years, we set $\tau = \frac{\ln(2)}{1.5 \cdot 365}$. Other parameters are $\sigma = 1/T_{incubation} = 1/4.6$ and $\gamma = 1/T_{infectious} = 1/2.1$. The relationship between these numbers and $R_0$ is as follows

$$R_0 = (1-\theta)\beta T_{infectious} \rho(\text{diag}(\mathbf{w})\mathbf{A}), \tag{7}$$

where $\rho$ denotes the spectral radius.

We start our modeling on the first of September when Stockholm county already had around 80 new cases per day. To get the distribution within the respective sub-groups right, we run the model with $\tau = 0$, starting from 1 exposed, until this number of new infections is hit. We then use these numbers as initial values in the compartments $\mathbf{e}$ and $\mathbf{i}$. Since $\mathbf{r}$ on September first already contain 10% with anti-bodies, we set $\mathbf{r} = 0.1\mathbf{w}$ and initialize $\mathbf{s}$ by setting $\mathbf{s}(0) = \mathbf{w} - \mathbf{e}(0) - \mathbf{i}(0) - \mathbf{r}(0)$.

To include variants of concern, we introduce new parameters $\beta_{voc}$ and $\theta_{voc}$ as well as functions $\mathbf{e}_{voc}$ and $\mathbf{i}_{voc}$, set

$$\boldsymbol{\nu}_{voc}(t) = \beta_{voc} \text{diag}(\mathbf{s}(t) - \theta_{voc} \mathbf{w}) A \mathbf{i}_{voc}(t), \tag{8}$$

and obtain an equation system where both original strain and voc's exist in parallel as follows:

$$\begin{cases} \mathbf{s}'(t) = -\boldsymbol{\nu}(t) - \boldsymbol{\nu}_{voc}(t) + \tau \mathbf{r}(t) \\ \mathbf{e}'(t) = \boldsymbol{\nu}(t) - \sigma \mathbf{e}(t) \\ \mathbf{e}'_{voc}(t) = \boldsymbol{\nu}_{voc}(t) - \sigma \mathbf{e}_{voc}(t) \\ \mathbf{i}'(t) = \sigma \mathbf{e}(t) - \gamma \mathbf{i}(t) \\ \mathbf{i}'_{voc}(t) = \sigma \mathbf{e}_{voc}(t) - \gamma \mathbf{i}_{voc}(t) \\ \mathbf{r}'(t) = \gamma \mathbf{i}(t) + \gamma \mathbf{i}_{voc}(t) - \tau \mathbf{r}(t) \end{cases}$$

#### 5.2 Calibration of the contact matrix

According to Public Health Agency data, 11.5% of the COVID-19 cases are from the age group 0-19, 35.5% in the age group 20-39, 43.5% in the age group 40-69 and 9.5% of the cases are in the group 70+ (from Feb. 2, before the effect of vaccinations kicked in). However, the model described above is based on a contact pattern matrix [26] which was measured before the COVID-19 pandemic. In Sweden, as a consequence of the persistently high society spread, the group 70+ have to a large extent self-isolated or at least kept their non-essential contacts to a minimum. Another issue is that the number of cases in the group 0-19 is misleading since testing for children has been severely limited. Based on blood donor data from the same period, it is estimated that 25% of the cases were in this group, which is also consistent with data from neighboring countries in the range 20-25%. We therefore added 12.5% to the age-group 0-19 and subtracted corresponding fractions from the remaining groups, leading to the ratios $(24, 31, 37, 8)$.

In [6] it is described how to take isolation of the 70+ group into account by modifying the last row and column of the contact matrix by a reduction factor $\xi$. To find a realistic value of $\xi$, we ran the above model for various values of $\xi$, and compared the outcome in the age-groups of the model with the numbers stated above. The reduction factor $\xi = 1/4$ gives values 23%, 32%, 37% and 8%, which thus gives support for this choice and moreover in general validates the use of the contact matrix from [26] in a Swedish setting.

#### 5.3 The herd immunity threshold

In this section we discuss how to estimate when herd-immunity is reached, given the model (5)-(6), i.e. we consider models involving multiple strains. This is because we are primarily interested in determining when the HIT occurred, and in the case of Stockholm, both times we had only one dominant strain. Even under this simplification, it is not entirely obvious of how to define the herd-immunity threshold, given that we work with multi-compartment models (various age-groups).

Intuitively, herd-immunity is reached when the amount of susceptibles are few enough that the amount of people who become sick, i.e. the total in $\mathbf{e}$ and $(i)$, start to recede. This is equivalent to demand that if we have no spread and a group of infectives are added to the system, the amount of secondary cases will be a decreasing function. The issue becomes delicate since this does not only depend on the amount of remaining susceptibles, but also on their distribution between the various groups. Classical estimates of HIT, (see [3] Ch. 5), assume a uniform distribution of immunity, whereas during a real outbreak immunity will be higher in groups that are more active in spreading the virus.

To take a concrete example, the mathematical (homogenous) HIT is reached on day 115 of our modeling, i.e. 2 days before Christmas, but the amount of individuals in all subgroups of both $\mathbf{e}$ and $\mathbf{i}$ start to decrease on day 95, i.e. third of December, just before the first update in the NPI's that could have had a limiting effect on the spread (closure of high-schools, see Fig. 1).

We now describe mathematically how to arrive at these numbers. The equations for $\mathbf{e}'$ and $\mathbf{i}'$ in (5)-(6), written using standard matrix-vector notation, become

$$\begin{bmatrix} \mathbf{e}'(t) \\ \mathbf{i}'(t) \end{bmatrix} = \begin{bmatrix} \beta \mathrm{diag}(\mathbf{s}(t) - \theta\mathbf{w})A\mathbf{i}(t) - \sigma\mathbf{e}(t) \\ \sigma\mathbf{e}(t) - \gamma\mathbf{i}(t) \end{bmatrix} = \begin{bmatrix} -\sigma I & \beta \mathrm{diag}(\mathbf{s}(t) - \theta\mathbf{w})A \\ \sigma I & -\gamma I \end{bmatrix} \begin{bmatrix} \mathbf{e}(t) \\ \mathbf{i}(t) \end{bmatrix} \tag{9}$$

where $I$ denotes the identity matrix. Following [3], Ch. 5.2, we shall call the matrix on the left hand side for $F - V$, where $F$ only consists of zeros and $\beta \mathrm{diag}(\mathbf{s}(t) - \theta\mathbf{w})A$ in the upper left corner, and $V$ contain the identity matrices (with zero in the upper left corner). We prove the following theorem.

**Theorem 5.1.** *Assume that $A$ is a symmetric matrix. Then the eigenvalues of $F - V$ are real and negative if and only if*

$$\rho(\beta \mathrm{diag}(\mathbf{s}(t) - \theta\mathbf{w})A/\gamma) < 1, \tag{10}$$

*where $\rho$ denotes the spectral radius of a matrix.*

Note that the contact matrix $A$ in reality should be symmetric, as shown in Sec. 7.1 of [6]. Hence, the point at which all compartments in the disease states $\mathbf{e}$ and $\mathbf{i}$ start to decrease, equals the first value of $t$ for which (10) is fulfilled.

*Proof.* By Lemma 5.2 of [3] and some straightforward computations (see Section 7.5 of [6]), it follows that $F - V$ has negative real parts if and only if the spectral radius of $FV^{-1}$, denoted $\rho(FV^{-1})$, is less than 1. Further more, this in turn is equivalent with (5.3) (again, see Section 7.5 of [6] for the relatively simple computations leading to this conclusion.) It remains to prove that under this assumption, the eigenvalues are actually real.

Eigenvalues $\mathbf{x}$ of $\beta \mathrm{diag}(\mathbf{s}(t) - \theta\mathbf{w})A$ also solve the generalized eigenvalue problem

$$\alpha(\mathrm{diag}(\mathbf{s}(t) - \theta\mathbf{w}))^{-1}\mathbf{x} = \beta A\mathbf{x},$$

where $\alpha$ is the eigenvalue in question. By standard theory of such equation systems, there exists a basis of real eigenvectors to such equation systems, with corresponding real eigenvalues, see e.g. [21]. To finish the proof, we show that each such solution $(\alpha, \mathbf{x})$ gives rise to two real eigenvalues/eigenvectors of $(F-V)$. Indeed, if we consider a vector of twice the size given by $[a\mathbf{x}, b\mathbf{x}]^t$ (where $t$ denotes the transpose), it follows that this is an eigenvector if and only if $[a,b]^t$ is an eigenvector of

$$\begin{bmatrix} -\sigma & \alpha \\ \sigma & -\gamma \end{bmatrix}.$$

By this we already have that the basis of generalized eigenvectors considered earlier extends to a basis of twice the size for the space on which $F-V$ operates. Finally, the characteristic polynomial of the above matrix has solutions

$$\lambda_{1,2} = -\frac{\sigma+\gamma}{2} \pm \sqrt{\frac{(\sigma+\gamma)^2}{4} - \sigma\alpha}.$$

By (10) we have that $|\alpha| < \gamma$, so the expression under the root is bounded from below by

$$\frac{(\sigma+\gamma)^2}{4} - \sigma\gamma = \frac{(\sigma-\gamma)^2}{4} \geq 0.$$

Hence the roots are real, and the proof is complete.

□

We now derive the more standard way of defining the herd-immunity threshold. Instead of inserting $\mathbf{s}(t)$ in (10), where $\mathbf{s}$ is obtained by solving SEIR, one may replace $\mathbf{s}(t)$ with the initial value $\mathbf{s}(0)$ and ask what level of immunity is needed in order to achieve that $(F-V)$ becomes a matrix whose eigenvalues are negative and real. Since $\mathbf{w}$ is the fraction of people in each subgroup, it becomes natural to set $\mathbf{s}(0) = (1-\psi)\mathbf{w}$, where $\psi$ now denotes the level of acquired immunity at time 0, and $\theta$ the level of (mathematical) pre-immunity. This leads to the equation

$$\rho(\beta \mathrm{diag}((1-\psi-\theta)\mathbf{w})A/\gamma) < 1,$$

which, upon recalling (7), can be recast in the following more attractive form

$$\psi = (1-\theta)(1-\frac{1}{R_0}). \tag{11}$$

The effect of population pre-immunity is thus to reduce the herd-immunity threshold by the corresponding fraction, which is why this threshold is in reality much lower than traditional models predict (i.e. upon setting $\theta = 0$).

For example, given the parameters used in this paper for the second wave of Stockholm, the herd-immunity threshold as given by (11) is $17.75\%$. This amount of acquired immunity is reached on day 115 of our model, whereas the equation (10) is satisfied from day 95 and onwards, when all disease compartments de facto start to shrink. In summary, we argue that (10) is the more realistic definition of the herd-immunity threshold in a concrete modeling example, whereas (11) is a good first order approximation (upper bound). In practice, our modeling experience suggests the two definitions are off by a couple of weeks at the most.

### 5.4 Incorporating vaccinations

We accessed and interpolated the curve of total fraction of the population vaccinated in Stockholm County, as a function $v(t)$, from the Public Health Agency [12]. The degree of protection from vaccinations will undoubtedly increase over time in the population, but since vaccine efficacy varies between different vaccines and in different age groups, it is not entirely straight forward how to include vaccines in a model where immunity is a binary value. The Astra Zeneca vaccine was given to elderly people and to health care personnel in the beginning of 2020, whereafter the mRNA vaccines from Pfizer and Moderna became the major vaccines given to all age groups in Sweden. While the Moderna and Pfizer vaccines are expected to have a protection above 90%, the Astra Zeneca vaccine is estimated at a 70% protective effect to the original SARS-CoV-2 strain, a reduced protection rate is expected against the alpha and delta variants. The alpha strain caused the third wave in Stockholm, and the delta strain is now slowly emerging. To model the vaccine protection, we add an "artificial" flow from the group $s$ to the group $r$, which now represent "recovered and vaccinated". Since we conclude that vaccination had a limited effect on the development in Stockholm, we prefer to use a simplified model where vaccine efficacy is exaggerated. Therefore, we postulate that $90\%$ of vaccinated people become "fully protected" (i.e. are moved from $s$ to $r$) three weeks after receiving their first shot. In our model, the highest age-group $70+$ make up $15\%$ (this is group number 6), which means that younger age groups were not beginning vaccination until early April. Another thing to keep in mind is that the vaccination scheme did not separate between people who had had COVID-19 and those who had not. Therefore, many people that got vaccinated were already in (the corresponding age-group of) the $r$-compartment. Taking these factors into account, we propose to model the vaccination roll-out by subtracting the term $0.9v'(t-21)s_6(t)/w_6$ to the last row of the equations for $s'$, and simultaneously add it to the last row of the $r'$ equations, where $t = 0$ is set at New Years eve 2020. Note that $s_6(t)/w_6$ gives the fraction of people in group 6 who are still susceptible at time $t$, which reflects the fact that many who were vaccinated already belonged to the group $r$. We do this as long as $v(t-21)$ stays below $0.8 \cdot 15\%$ (80% is the maximum achieved vaccination coverage in any group achieved up until end of May). When this point is reached, the we change the equation system so that the vaccinations affect group 5, i.e. those aged 40-69 who constitute $39\%$. Since our modeling stops early June, there is no need to move to group 4.

### 5.5 Modeling with variable transmission rate $\beta$.

The transmission rate $\beta$ will vary over time partially due to the introduction of VoC's, and partially due to changes in peoples behavior and NPI's. To keep these separate, we replace the constant $\beta$ with the product of two functions $\beta(t)$ and $f(t)$, where the latter only model changes in behavior/NPI. The former will be constant until the arrival of VoC's in early January, by which time it will start to grow and reach a new level once the VoC's have taken over, in early April, (see Figure 3). Since, due to Figure (3), we know the fraction of VoC's over time, we can form $\beta(t)$ easily by setting

$$\beta(t) = \beta_{OS}\frac{Cases_{OS}(t)}{Cases_{Tot}(t)} + \beta_{VoC}\frac{Cases_{VoC}(t)}{Cases_{Tot}(t)}$$

where $Cases_{OS}$, $Cases_{VoC}$ and $Cases_{Total}$ represent the red, yellow and blue curve in Figure 3 respectively. The constants $\beta_{OS}$ and $\beta_{VoC}$, representing the transmission rate of Original Strain and the VoC's, will be determined shortly.

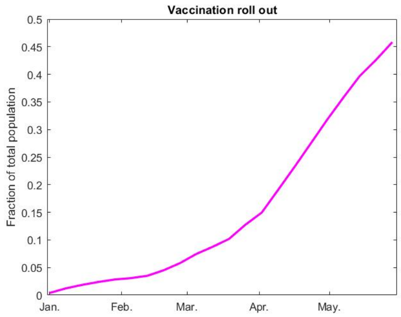


Figure 6: Fraction vaccinated $v$ as function of time $t$.

To construct a new model, we reuse the equation system (6), with the new feature that (5) is replaced by

$$\boldsymbol{\nu}(t) = f(t)\beta(t)\mathrm{diag}(\mathbf{s}(t))A\mathbf{i}(t), \tag{12}$$

where $f(t)$ on each day is chosen so that the total amount of new cases coincides with $Cases_{Tot}$ (blue curve in Figure 3-4). We therefore get a perfect model fit and there is no need to display graphs of the type 4.

As mentioned in 4.2, we pick the choice of the original transmission rate $\beta_{OS}$ so that the average of $f$ in September equals one. Once a value of $\beta_{OS}$ has been fixed, the next question becomes how much higher to put $\beta_{VoC}$. As argued in the main text, data seem to support a value around $56\%$, but we have chosen $\beta_{VoC} = 1.38\beta_{OS}$ in order to favor low values of $f$.
